# Antenatal magnesium sulphate for neuroprotection of preterm babies: data analysis of inequities in treatment in England 2014 to 2024

**DOI:** 10.64898/2026.01.06.26343507

**Authors:** Hannah B Edwards, Lauren J Scott, Cheryl McQuire, Isobel L Ward, David Odd, Frank de Vocht, Karen Luyt

**Author notes:** Corresponding author: Hannah B Edwards, Address: NIHR ARC West, Level 9 Whitefriars, Lewins Mead, Bristol BS12NT.

## Abstract

**Objectives:** To identify (a) sociodemographic inequities in treatment with antenatal MgSO_4_ for fetal neuroprotection in preterm birth, and (b) whether any inequities have improved following implementation of a national quality improvement programme to improve use of MgSO_4_.

**Design:** Secondary data analysis study using adjusted logistic regression modelling.

**Setting:** NHS England maternity units from 2014 to 2024.

**Participants:** Mothers with a preterm baby born from 24 to under 30 weeks’ gestation and admitted to an NHS neonatal unit.

**Outcomes:** Receipt of antenatal MgSO_4_.

**Results:** Before implementation of the National *PReCePT* (Preventing Cerebral Palsy in Pre-Term labour) Programme (NPP), mothers from the North of England had lower odds of MgSO_4_ treatment than mothers from the South of England (OR 0.60, 95% CI 0.49 to 0.74, p<0.001). Following implementation of the NPP, there was no longer evidence for a regional inequity in treatment (OR 0.87, 95% CI 0.70 to 1.08, p=0.201). Pre-NPP, mothers from more deprived areas had lower odds of treatment than women from more affluent areas (OR 0.87, 95% CI 0.80 to 0.95, p=0.002), but again, post-NPP, there was no longer evidence for inequity by deprivation (OR 1.02, 95% CI 0.92 to 1.14, p=0.719, p-value for interaction=0.022). White British mothers had slightly lower odds of treatment than mothers of other ethnic groups (OR 0.93, 95% CI 0.87 to 1.00, p=0.038), and this inequity did not appear to change from pre to post-NPP (p-value for interaction=0.235). There was little evidence that maternal age affected odds of treatment.

**Conclusions:** Inequities in delivery of antenatal MgSO_4_ treatment for preterm birth have improved in England, following implementation of the National *PReCePT* quality improvement programme.

**STRENGTHS AND LIMITATIONS OF THIS STUDY:** Strengths:

- Study uses standardised, routinely, prospectively collected healthcare data from all NHS neonatal units in England reporting on about 40,000 preterm births.
- Data covered a 10-year period (2014 to 2024) and the intervention was implemented in 2018, which gives sufficient time for reliable trends to be observed.
- Statistical modelling estimated the total effects of each risk factor separately, adjusted for measured confounders, and models were developed *a priori* using Directed Acyclic Graphs.

Limitations:

- The dataset only includes data on live-born infants (and their mothers) who were admitted to a neonatal unit.
- There are other individual characteristics that could also affect equity of care that were not measured in this data and so could not be evaluated here.

**What is already known on this topic:**

- Uptake of antenatal magnesium sulphate (MgSO_4_), a neuroprotective treatment to protect against cerebral palsy in preterm births, has recently improved in England.
- This is at least in part due to implementation of a national quality improvement programme (*PReCePT*) designed to improve use of MgSO_4_.
- But there are persistent inequities in many aspects of maternity and perinatal care in England, and overall improvements in care do not imply improvements in equity.

**What this study adds:**

- Before implementation of *PReCePT*, mothers from the North of England, from more socioeconomically deprived areas, or were white British ethnicity, all had lower odds of receiving antenatal MgSO_4_ treatment.
- After implementation of *PReCePT*, antenatal MgSO_4_ treatment became more equitable with no statistically significant differences in MgSO_4_ treatment by region or socioeconomic status.

**How this study might affect research, practice or policy:**

- Perinatal optimisation programmes should formally evaluate and aim to improve equity of care.

## INTRODUCTION

Persistent inequities in maternity and perinatal care and outcomes are well documented in the UK. The *Royal College of Obstetricians and Gynaecologists* National Maternity and Perinatal Audit (2021) reported ethnic and socioeconomic differences in caesarean birth, postpartum blood loss, small-for-gestational-age (SGA) births, Apgar scores, neonatal admissions, and breastfeeding rates.(1) *MBRRACE-UK* Confidential Enquiries have consistently identified ethnic inequities in the care of women who experienced stillbirth, neonatal death, or maternal death.(2) The *National Neonatal Audit Programme* has highlighted geographical variation in antenatal preterm optimisation interventions, deferred cord clamping, neonatal mortality, and key morbidities such as bronchopulmonary dysplasia and necrotising enterocolitis.(3) The 2025 report from the National Child Mortality Database showed clear patterning of child deaths by ethnicity and deprivation, with perinatal or neonatal events as the leading cause of death.(4) Recent UK cohort studies confirm continuing ethnic, socioeconomic, and maternal age-related inequities in stillbirth, preterm birth, fetal growth restriction, and neonatal mortality.(5–7) Comparable patterns are seen in most other high– and upper-middle-income countries.(8–10)

Reducing these inequities is a stated priority for the UK government, which has recently established a national maternity and neonatal taskforce and committed to an independent investigation of maternity and neonatal services(11, 12)

Administration of antenatal magnesium sulphate (MgSO□) to women in preterm labour is a key evidence-based intervention recommended by both the National Institute for Health and Care Excellence (NICE)(13) and the World Health Organisation (WHO).(14) Systematic review and meta-analysis demonstrate that MgSO_4_ reduces the risk of cerebral palsy (CP) in preterm infants by around 30%.(15) Preventing CP is of major importance given its lifelong effects on motor, cognitive, and sensory function, and its substantial personal and economic costs.(16, 17) Avoidable cases of CP account for half of NHS litigation costs, with average payouts exceeding £10 million per case.(18) MgSO□ is inexpensive and highly cost-effective, generating an estimated £1 million in lifetime societal savings per case prevented.(19, 20)

Uptake of antenatal MgSO_4_ in the UK has improved following the National *PReCePT* (Preventing Cerebral Palsy in Pre-Term labour) quality improvement (QI) programme.(21, 22) *PReCePT* provided maternity units with clinical guidance, staff training, educational resources, and midwife champions to lead local delivery.(23, 24) However, overall healthcare quality improvements can mask, or even worsen underlying inequities in care.(10, 25–29) It is uncommon for QI interventions to properly evaluate, let alone address inequities.(30–32) Given the pervasive inequities across other aspects of maternity and perinatal care, similar patterns in uptake of antenatal MgSO_4_ are plausible. They are also crucial to address given that inequity in preventative treatment here will perpetuate costly, lifelong health and social inequities.

## AIMS

This study aimed to identify (a) sociodemographic inequities in treatment with antenatal MgSOD, and (b) whether any inequities have improved following implementation of a national quality improvement programme (the National *PReCePT* Programme (NPP)) to improve use of MgSO□. Maternal age, socioeconomic deprivation, ethnicity, and geographical region were the key factors of interest. Our hypothesis was that MgSO_4_ administration had become more equitable following implementation of the NPP.

## METHODS

### Design

This was a secondary data analysis study.

### Study setting and population

The study setting was England, January 2014 to December 2024. The population was mothers of babies born preterm between 24 weeks and zero days gestation (24^+0^), and 29 weeks and six days gestation (29^+6^) (inclusive). Data were extracted from the National Neonatal Research Database (NNRD), which holds data on all live-born preterm babies that were admitted to an NHS neonatal unit in England.(33)

### Outcomes and exposures

The outcome was receipt of MgSO_4_. Exposures of interest were maternal age (≥35 versus <35 years, as age 35 and over is the definition of a ‘geriatric’ pregnancy); area deprivation (Index of Multiple Deprivation decile,(34) coded as the five most versus the five least deprived deciles); ethnicity (white British versus all other groups, based on mother’s declared ethnicity from the NHS England standard codes for ethnic group); and region (North versus South of England, coded by regional neonatal Operational Delivery Network (ODN)). The time periods of interest (pre-versus post-NPP) was defined at the hospital level, as although the NPP was formally rolled-out in May 2018, individual hospitals varied in their start dates (from April 2018 to October 2020). Further details of variable coding are described in the study protocol.(35)

Covariates considered for model inclusion were smoking history (any record versus none); single versus multiple birth; number of previous pregnancies (none versus one or more); fever in labour; receipt of antibiotics in labour; premature rupture of membranes (PROM); meconium stained liquor; antenatal steroids given; gestational age (GA; continuous completed weeks and days); onset of labour (spontaneous preterm labour or not); mode of delivery (vaginal versus caesarean). Covariate selection was informed by Directed Acyclic Graphs (DAGs), developed *a priori* by the research team.(36–38) Appendices 1-3 present the DAGs and minimally sufficient adjustment sets for each exposure of interest. Further details are available in the study protocol.(35)

### Analysis

Categorical data are presented as numbers and percentages, and continuous data are presented as means and standard deviations (SD), or medians and interquartile ranges (IQR) if not normally distributed. Logistic regression models with the outcome being receipt of MgSO_4_ were developed for each of the four sociodemographic factors separately (maternal age, deprivation, ethnicity, and region), generating Odds Ratios (OR) for receiving MgSO_4_ associated with that sociodemographic factor. Variables in the unadjusted models included the factor of interest, NPP period, and interaction between sociodemographic factor and NPP period. This interaction term was included to identify changes in associations from pre– to post-NPP. Models were also run without the interaction term to obtain the overall associations.

An adjusted model was also developed for each sociodemographic factor separately to estimate the *total effects* of each sociodemographic factor on the outcome. All of these additionally adjusted for date (calendar month and year), and clustering at the level of the neonatal unit, as mothers attending the same unit are likely to experience more similar care compared to mothers attending different units.

Due to antenatal MgSO_4_ also having a therapeutic role in the management of pregnancy hypertension and pre-eclampsia, models were re-run excluding mothers with pregnancy hypertension as a sensitivity analysis.

As a further supplementary analysis, we also developed adjusted models to estimate the *direct effects* of each sociodemographic factor on the outcome (the direct effects representing the component of total effects not mediated by indirect causal pathways(39), Appendices 1-4). All models were specified *a priori* based on DAGs (Appendices 1-3 and study protocol(35)).

### Ethics

This research was granted Health Research Authority (HRA) approval (IRAS id: 260504) and institutional Faculty Research Ethics Committee (FREC) approval (FREC id: 5670).

### Patient and Public Involvement

Patients and the public were involved in the conduct, reporting, and dissemination of the wider programme of work that this study is part of; for example, two mothers with experience of preterm birth were on the programme Steering Committee.

## RESULTS

### Population description

Mother and baby characteristics were largely comparable pre– and post-NPP (Table 1). There were potential areas of difference in reported ethnicity (51.8% white British pre-NPP, 45.1% post-NPP); smoking history (17.3% pre-NPP, 14.1% post-NPP); meconium-stained liquor (5.2% pre-NPP, 1.9% post-NPP); receipt of antibiotics in labour (27.5% pre-NPP, 16.5% post-NPP); spontaneous preterm labour (57.2% pre-NPP, 48.3% post-NPP); and mode of delivery (55.2% caesarean section pre-NPP, 63.6% post-NPP). MgSO_4_ uptake improved from a mean 48.3% across the pre-NPP period, to a mean 86.5% across the post-NPP period.

**Table 1:** Characteristics of preterm births (24^+0^ to 29^+6^ weeks gestation) in England 2014-2024.

|  | **Pre National PReCePT Programme (2014-2018\*)** | **Post National PReCePT Programme (2018-2024\*)** |
| --- | --- | --- |
| **Number of mothers** | 16,625 | 18,377 |
| **Number of babies** | 19,076 | 20,698 |
| **Maternal age (mean, standard deviation (sd))** | 30.5 (6.2) | 31.2 (6.1) |
| Missing | 102 | 120 |
| **Maternal ethnicity (n, %)** |  |  |
| White British | 8,607 (51.8%) | 8,292 (45.1%) |
| All other groups | 5,421 (32.6%) | 6,071 (33.0%) |
| Missing | 2,597 (15.6%) | 4,014 (21.8%) |
| **Maternal Index of Multiple Deprivation (IMD) (median, inter-quartile range, (IQR))** | 4 (2, 7) | 4 (2, 7) |
| Missing | 301 | 266 |
| **Region** |  |  |
| North of England | 8,055 (48.5%) | 8,883 (48.3%) |
| South of England | 8,570 (51.5%) | 9,494 (51.7%) |
| **Smoking history (n, %)** |  |  |
| Yes | 2,874 (17.3%) | 2,593 (14.1%) |
| No | 11,761 (70.7%) | 13,116 (71.4%) |
| Missing | 1,990 (12.0%) | 2,668 (14.5%) |
| **Multiple birth (n, %)** | 2,336 (14.1%) | 2,208 (12.0%) |
| **Number of previous pregnancies (n, %)** |  |  |
| 0 | 5,516 (33.2%) | 6,292 (34.2%) |
| >0 | 9,984 (60.1%) | 10,386 (56.5%) |
| Missing | 1,125 (6.8%) | 1,699 (9.2%) |
| **Fever in labour (n, %)** | 951 (5.7%) | 705 (3.8%) |
| **Received antibiotics in labour (n, %)** | 4,564 (27.5%) | 3,026 (16.5%) |
| **Premature rupture of membranes (PROM) (n, %)** | 2,699 (16.2%) | 3,095 (16.8%) |
| **Meconium-stained liquor (n, %)** | 868 (5.2%) | 345 (1.9%) |
| **Gestational age (weeks, median, IQR)** | 27.9 (26.3, 29.0) | 27.7 (26.3, 28.9) |
| **Spontaneous preterm labour (n, %)** |  |  |
| Yes | 9,512 (57.2%) | 8,882 (48.3%) |
| No | 6,214 (37.4%) | 8,202 (44.6%) |
| Missing | 899 (5.4%) | 1,293 (7.0%) |
| **Mode of delivery (n, %)** |  |  |
| Vaginal | 7,451 (44.8%) | 6,697 (36.4%) |
| Caesarean | 9,174 (55.2%) | 11,680 (63.6%) |
| **Antenatal steroids given (n, %)** |  |  |
| Yes | 15,313 (92.1%) | 17,241 (93.8%) |
| No | 1,234 (7.4%) | 1,110 (6.0%) |
| Missing | 78 (0.5%) | 26 (0.1%) |
| **Antenatal magnesium sulphate given (n, %)** |  |  |
| Yes | 8,022 (48.3%) | 15,902 (86.5%) |
| No | 5,643 (33.9%) | 2,397 (13.0%) |
| Missing | 2,960 (17.8%) | 78 (0.4%) |

\*May 2018 was the NPP national launch date, but individual hospitals varied in their start dates. Figures presented are indexed on individual hospital start dates.

### Maternal age

There was little evidence that maternal age was associated with MgSO_4_ delivery (total effects Adjusted OR (aOR) 1.01, 95% CI 0.94, 1.08, p=0.839, with comparable results in the unadjusted model) (Table 2). There was also little evidence for a change in this association from pre– to post-NPP, although coefficients suggested that older mothers may have had slightly higher odds of treatment pre-NPP, that equalised post-NPP (Table 2, Figure 1a).

**Figure 1a:**
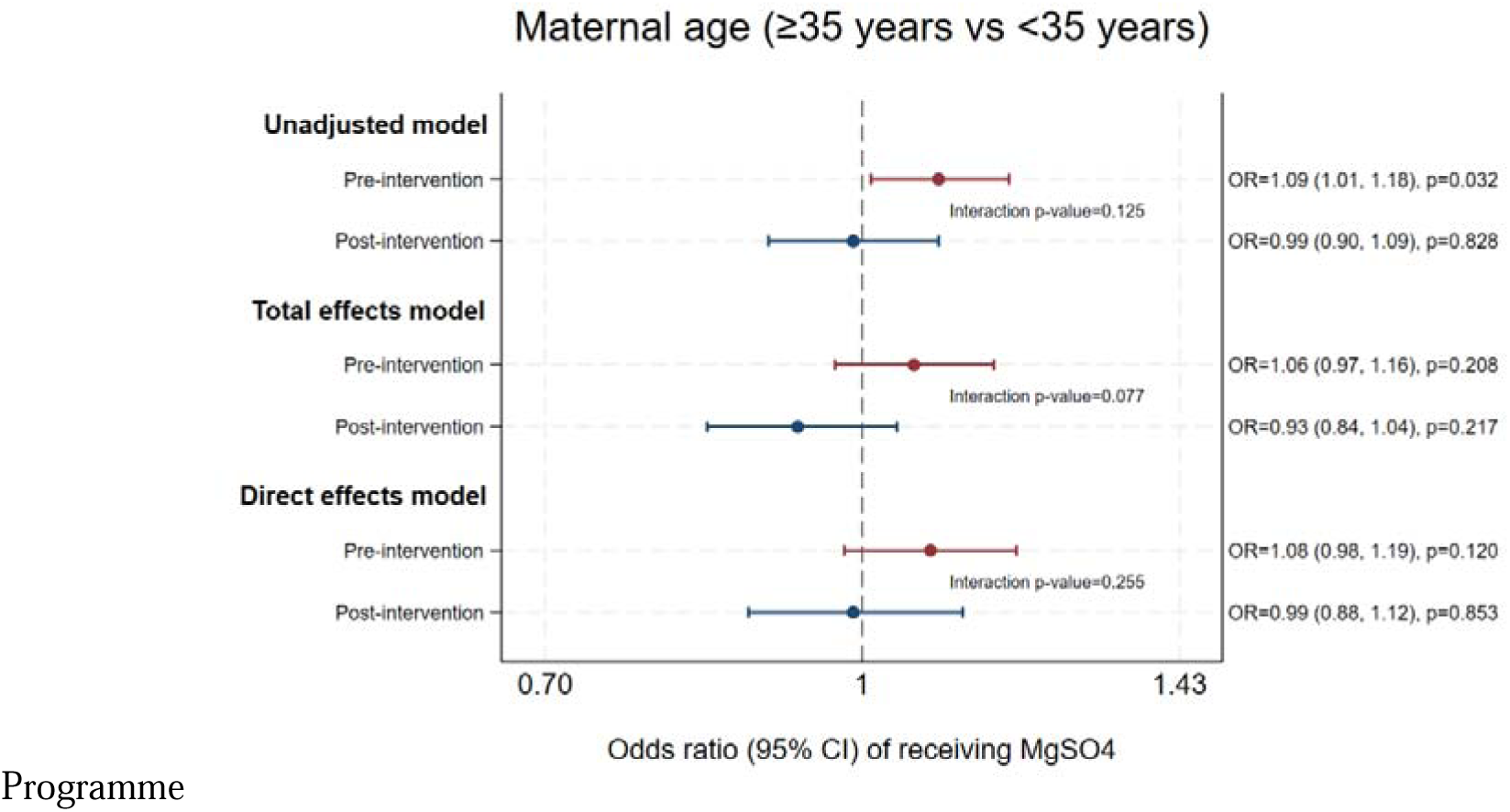
The effect of maternal age on odds of antenatal MgSO4 treatment, pre and post National PReCePT Programme.

**Table 2:**
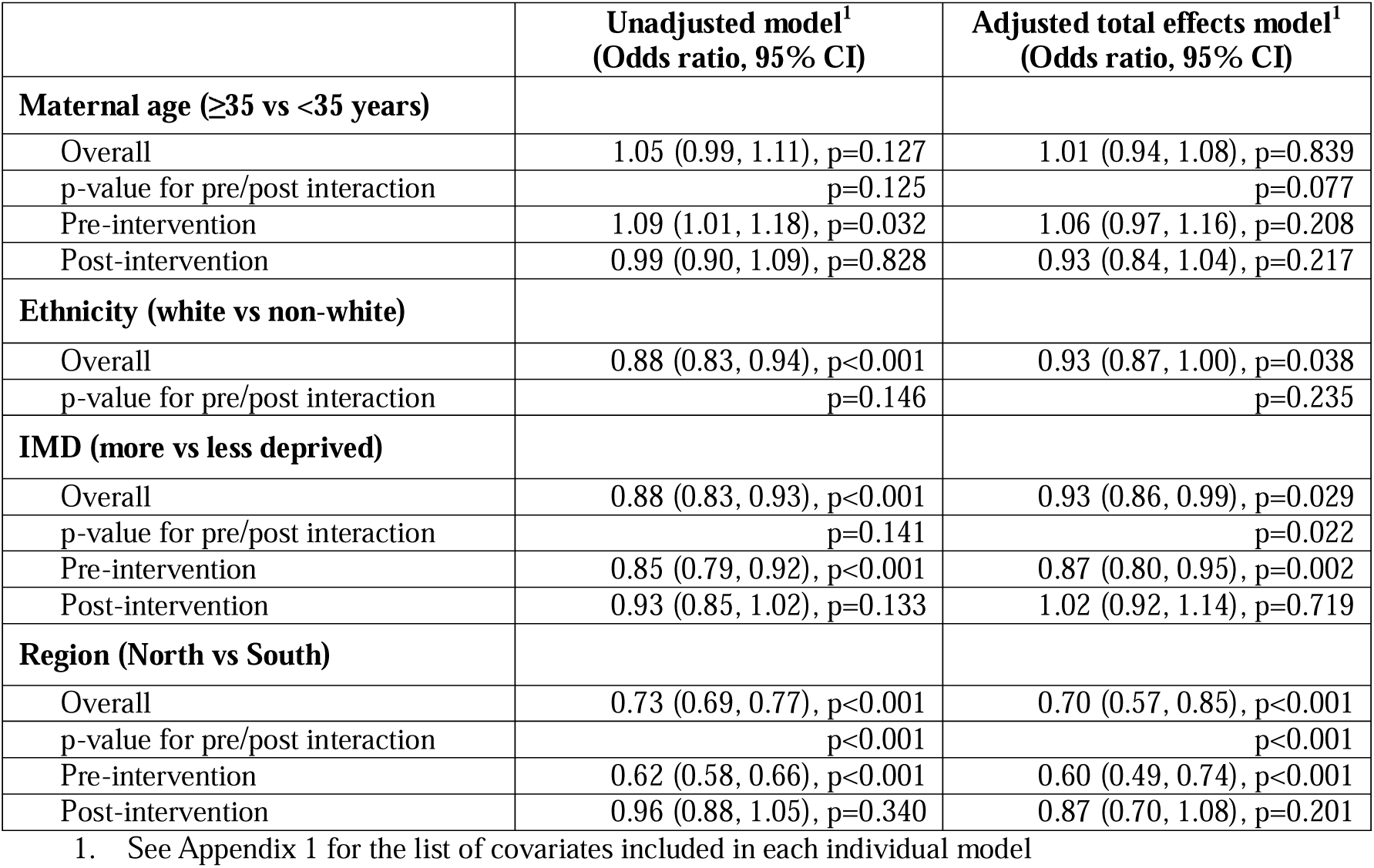
Total effects of sociodemographic factors on odds of receiving antenatal magnesium sulphate for preterm birth in England 2014-2024.

### Ethnicity

The unadjusted model showed that mothers of white British ethnicity had lower overall odds of treatment compared to mothers of other ethnicities, a difference that attenuated slightly after adjustment (aOR 0.93, 95% CI 0.87, 1.00, p=0.038) (Table 2). There was no evidence that this inequity changed from pre– to post-NPP (Table 2, Figure 1b).

**Figure 1b:**
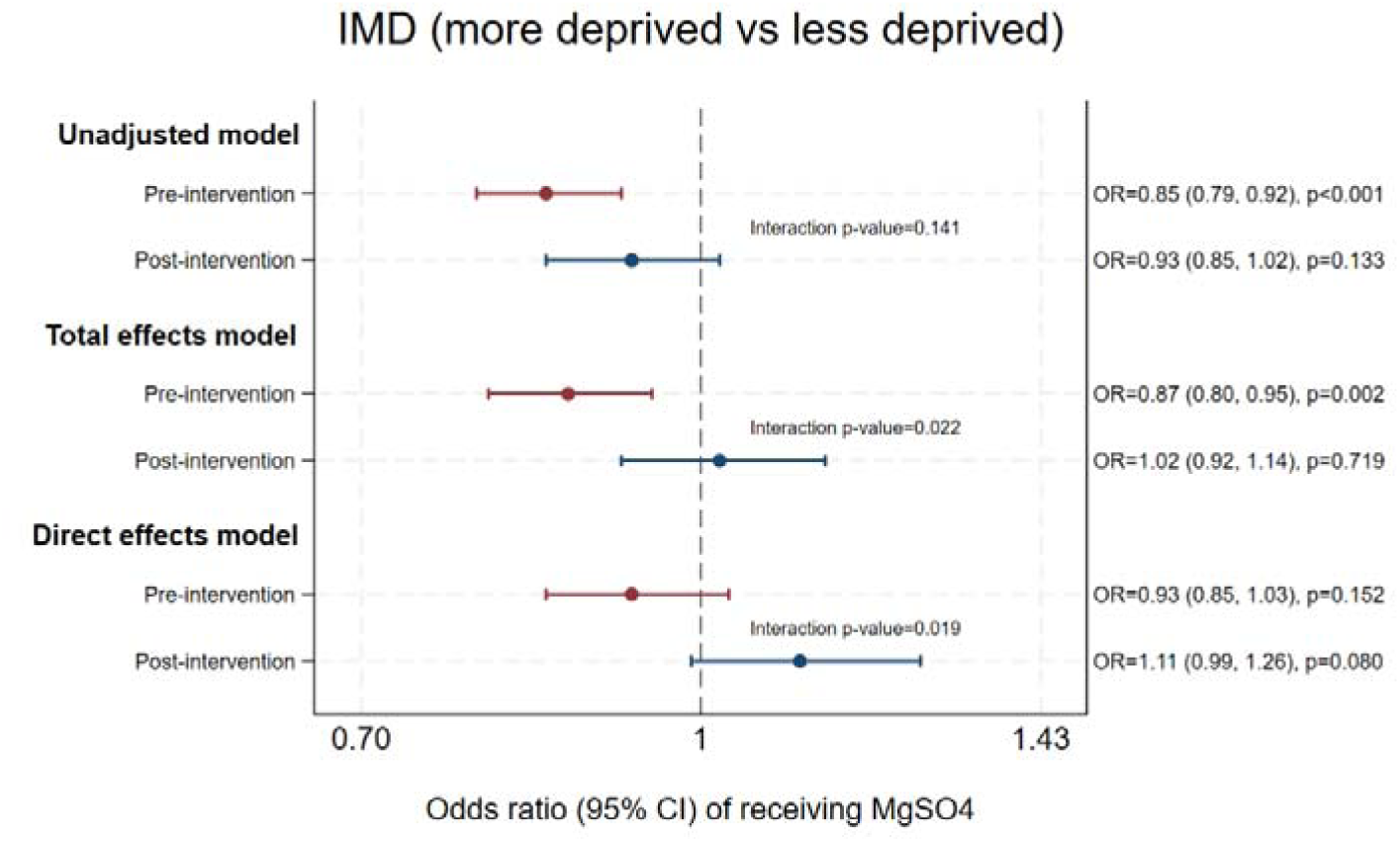
The effect of area deprivation on odds of antenatal MgSO4 treatment, pre and post National PReCePT Programme.

### Area deprivation

The unadjusted model showed that mothers from more deprived areas had lower overall odds of MgSO_4_ treatment compared to mothers from less deprived areas, a difference that attenuated slightly after adjustment (aOR 0.93, 95% CI 0.86, 0.99, p=0.029) (Table 2). There was evidence for an improvement in this inequity from pre– to post-NPP (interaction p=0.022): pre-NPP, mothers from more deprived areas had lower odds of treatment (aOR 0.87, 95% CI 0.80, 0.95, p=0.002); post-NPP, there was no longer evidence that deprivation level affected odds of treatment (OR 1.02, 95% CI 0.92, 1.14, p=0.719) (Table 2, Figure 1c).

**Figure 1c:**
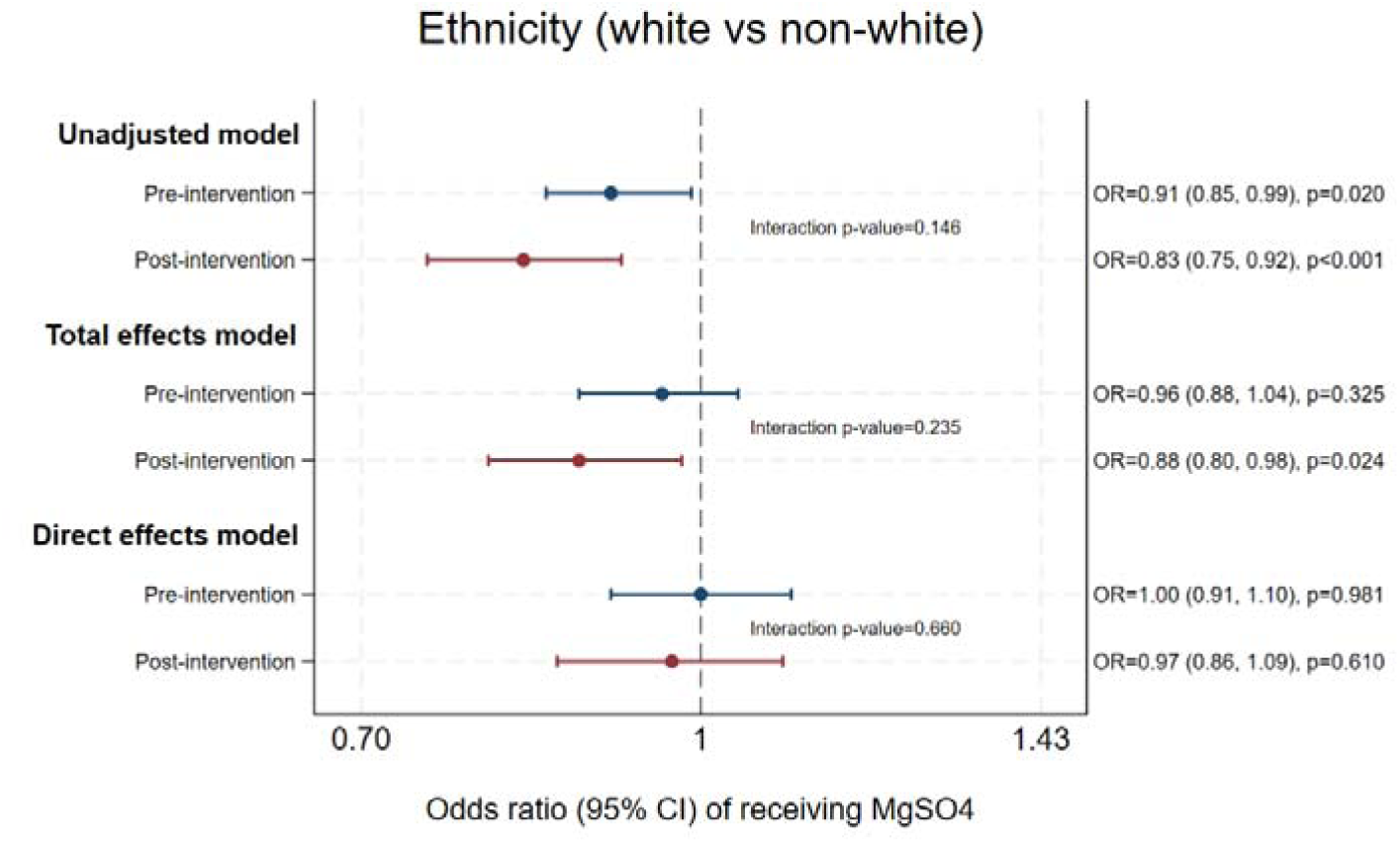
The effect of maternal ethnicity on odds of antenatal MgSO4 treatment, pre and post National PReCePT Programme.

### Region

There was strong evidence for a significant regional inequity in MgSO_4_ treatment: pre-NPP, mothers in the North of England had around 40% lower odds of treatment compared to mothers in the South of England (total effects aOR 0.60, 95% CI 0.49, 0.74, p<0.001) (Table 2). Post-NPP, there was no longer evidence that region affected odds of treatment (total effects aOR 0.87, 95% CI 0.70, 1.08, p=0.201) (Table 2, Figure 1d).

**Figure 1d:**
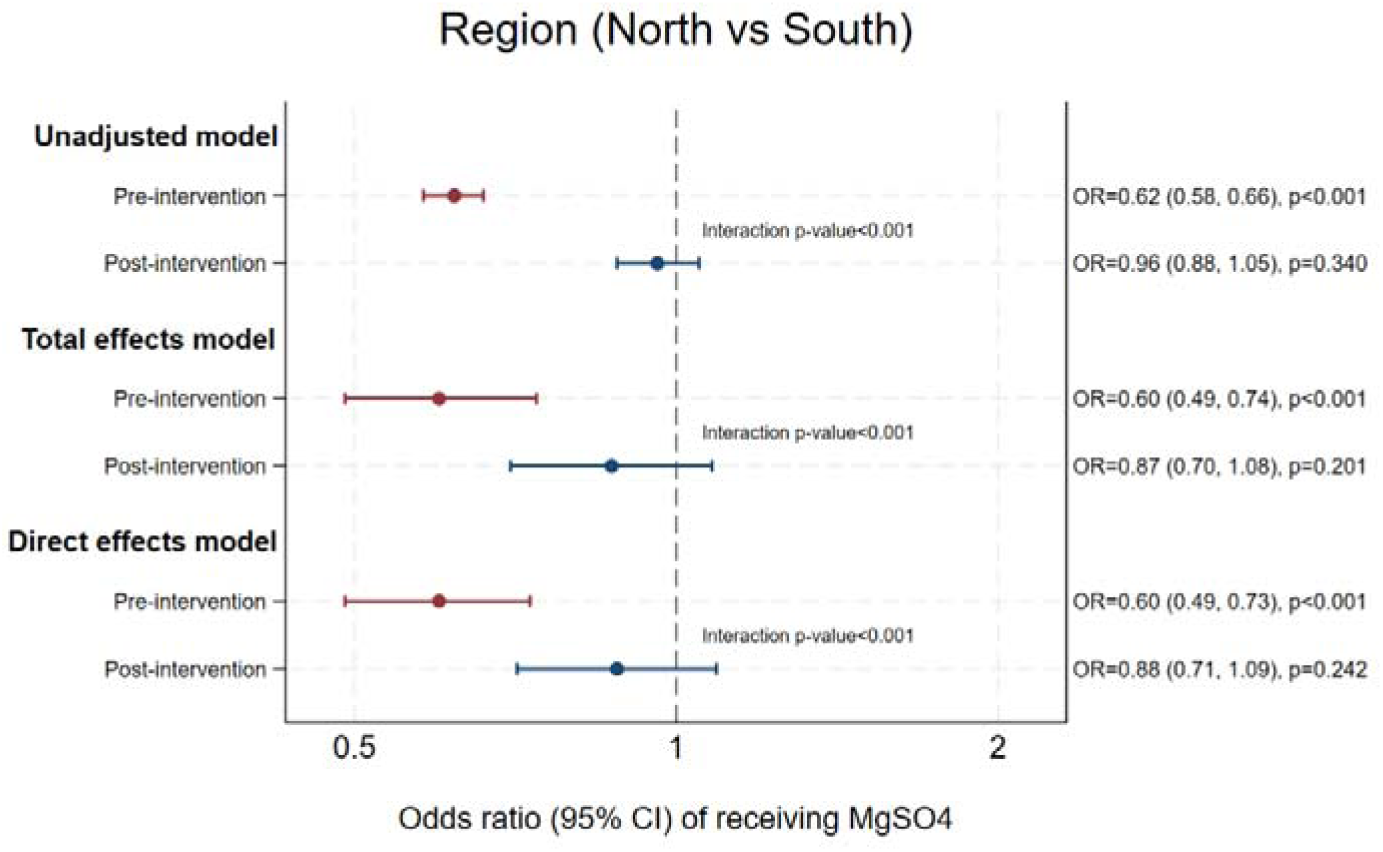
The effect of region (North versus South of England) on odds of antenatal MgSO4 treatment, pre and post National PReCePT Programme.

### Supplementary analyses

Statistical models adjusted for the *direct effects* of each sociodemographic factor (i.e., excluding effects via indirect causal pathways) indicated that there was little evidence for a direct effect of maternal age, ethnicity, or deprivation on odds of treatment. There did appear to be a direct effect of region, with estimates very comparable to those found in the total effects model (Appendix 4).

### Sensitivity analyses

Excluding mothers with a record of pregnancy hypertension produced results compatible with the main analyses (Appendices 5-6).

## DISCUSSION

### Key findings

By far the strongest observed inequity in antenatal MgSO_4_ treatment for neuroprotection of preterm babies was between mothers in the North versus South of England: mothers in the North were around half as likely to receive antenatal MgSO_4_ compared to mothers in the South prior to NPP implementation. This striking regional inequity appeared to improve post-NPP. Mothers from more deprived areas were less likely to receive treatment compared to mothers from more affluent areas prior to NPP implementation. This inequity also appeared to improve post-NPP. White British mothers were less likely to receive MgSO_4_ treatment compared to mothers of other ethnicities (including white mothers of non-British ethnicity), and this inequity appeared consistent throughout the study period. There was little evidence that maternal age-affected odds of MgSO_4_ treatment, although estimates suggested that pre-NPP, older mothers may have had slightly higher odds of treatment.

### Comparison with other literature

The stark evidence for a historical, high-magnitude treatment inequity by North/South of England is consistent with other evidence on broader health inequities by the “North/South divide”.(3, 40) Inequity in protective antenatal care for preterm babies is particularly significant, as it drives (and may compound) a lifetime of health and economic burden for affected individuals, families, and society.(41–43) This inequity persisted through statistical adjustment for known key confounding factors including deprivation, ethnicity, and smoking status, and other differences in population characteristics between the two periods. Although it is likely that there are other unmeasured differences not captured here (a key one being patient behaviour), it is reassuring that since implementation of the NPP, there is no longer evidence of this inequity.

Deprivation is well-known to have adverse effects on receipt of healthcare, and is believed to act through multiple entrenched mechanisms and at multiple levels.(44) Our findings that deprivation was associated with lower odds of MgSO_4_ treatment are consistent with this wider picture. How deprivation would drive differences in treatment in this context are not clear. The supplementary analysis found little evidence for a *direct* effect of deprivation on odds of MgSO_4_ treatment, indicating that this association is driven by indirect pathways. Further exploration of this would be valuable, but again it is reassuring that since implementation of the NPP, there is no longer evidence of this treatment inequity.

The slight disadvantage experienced by White British mothers in receipt of MgSO_4_ treatment is interesting, and counter to our hypothesis, as the direction of effect is opposite to that commonly found in other aspects of maternity and perinatal care. Commonly, black and ethnic minority groups are the ones at a disadvantage in both receipt of care and health outcomes.(1, 5–7, 45) However, comparable results of disadvantage to White British mothers in some aspects of maternity care have been found in the recent *MBRRACE* reports(2). Again, the supplementary analysis found little evidence for a *direct* effect of ethnicity on odds of MgSO_4_ treatment, indicating that this association is driven by indirect pathways (including, plausibly, factors not measured here). Further exploration of the relevant individual characteristics driving this association would be valuable. In particular, although difficult to measure, it would be interesting to further explore and untangle the relative contributions of cultural factors, race, and nationality.

The NPP was a universal intervention for English maternity units, and successfully improved overall treatment rates. Although overall improvement does not necessarily have any implications for equity, it is plausible that this intervention did have a role in the improvements in equity observed here. Mothers who were less likely to be treated pre-NPP may have been the ones who benefitted most from the intervention. This fits with the broader pattern of where there is a ceiling effect on uptake, groups with a lower starting uptake have the most room for improvement.

### Strengths and limitations

Key strengths of this study are that it uses standardised, routinely, prospectively collected healthcare data from all NHS neonatal units in England. This minimises biases in the data such as selection bias, observer bias, and recall bias. Findings are generalisable across the country. Data covered a 10-year period of 2014 to 2024, and the NPP intervention was implemented in 2018, which gives sufficient time for reliable trends to be observed. Statistical modelling estimated the total effects of each sociodemographic factor, adjusted for measured potential confounders, and as supplementary analyses we additionally estimated the direct effects of each of these key sociodemographic factors. All models were developed *a priori* using DAGs, to clarify assumptions and guide design of the statistical models to minimise confounding and best estimate causal effects. Results were robust to sensitivity analyses.

Observed differences in receipt of a treatment does not always indicate inequity in care. In some cases it could just reflect that different groups have different levels of need. However in this case, all mothers in this study are eligible for treatment, so are considered to have equal need. Medical contra-indication is rare and unlikely to be sociodemographically patterned. This indicates that the observed differences are likely to reflect a true inequity in care. Similarly, differences in treatment could be due to differences in patient behaviour – for example if certain patient groups tend to arrive later at hospital, so have a smaller window of opportunity for treatment. Arguably such cases are not ‘inequity in care’, or at least not inequity at the point of care, but just variations in patient behaviour that lead to differential treatment rates. Further exploration of upstream patient behaviour would be valuable here to better understand the causal pathways leading to differences in treatment. However, again we would argue that all of these mothers should be being treated, and if some groups are under-treated, that is an inequity to be addressed, regardless of whether the reason applies at the point of care or further upstream.

A limitation is that we did not have data on all possible covariates of interest, so residual confounding is plausible. For example maternal BMI, patterns and details of previous birth history (including assisted conception and stillbirths), how close the mother lives to their maternity unit, how early or late different groups of mothers tend to present at hospital, could all potentially affect receipt of MgSO_4_. Similarly, there are other individual characteristics that as well as being potential confounders, could also be considered important dimensions of equity themselves – e.g., race, language, sexual orientation, disability status, chronic illness status, marital status, education, health literacy, and urban/rural living(46) – that were not measured in this data and so could not be evaluated here.

It is possible that changes in population characteristics over time could be explaining some of the differences observed here. The post-NPP population tended to have slightly fewer smokers, slightly fewer cases of fever in labour or receiving antibiotics, fewer cases of meconium-stained liquor, more cases of spontaneous onset of labour, and more caesarean sections. It is plausible that some of these factors are also associated with likelihood of treatment – e.g. non-smokers may be more likely to be present and available to treat, and fewer ‘complications’ such as fever and meconium-stained liquor could mean a bigger window of opportunity to treat between presentation and delivery. We tried to address this by adjusting for these factors wherever indicated in the DAGs, but residual confounding is possible, and of course there may be alternative views on the details of some of the relationships modelled in our DAGs.

Another limitation is that the neonatal dataset only includes data on live-born infants who were admitted to a neonatal unit, and their mothers. This is a sub-group of an underlying population of mothers and infants who may also have been treated with MgSO_4_ for expected preterm birth, but who either did not go on to have a preterm birth, or where the infant died prior to NICU admission. If possible, it would be a valuable future research study to replicate this work on the wider underlying population.

More granular detail on individual-level deprivation factors may be useful in future work. It is possible that factors not captured by the area-level Index of Multiple Deprivation (IMD) (or captured, but different at the individual versus area-level) could also drive differences in receipt of healthcare. Similarly, more granular detail in the differences in treatment between the different minority ethnic groups would be valuable, as it is possible that combining them – as was indicated in this study to give sufficient power – may conceal variation in sub-group treatment rates.(10)

For the minority of eligible mothers not receiving antenatal MgSO_4_, further exploration of the reasons why they did not – and whether or not this varies by sociodemographic factors – would be valuable. Previous research on this population indicates that the overwhelming reported reason for not receiving treatment (>75% of cases) is ‘imminent delivery’, with smaller proportions reported as ‘not offered’ (11%), ‘not appropriate’ (5%), and very rarely ‘contraindicated’ or ‘declined’.(21) In this study the sample size would not have been sufficient to power further analysis of this by sociodemographic sub-groups, but if numbers allowed in larger, international datasets (for example the Vermont Oxford Network dataset(47)), further analysis on this question would be useful.

## CONCLUSIONS

Delivery of MgSO_4_ was patterned by deprivation, ethnicity, and possibly maternal age, as well as showing wide regional variation. Since the roll-out of *PReCePT*, a national QI programme that increased overall use of MgSO_4_, these inequities in care have improved, although White British mothers still have slightly lower odds of treatment than those of other ethnicities. This study illustrates that a nationally implemented QI programme can also improve equity of care, and highlights the importance of evaluating for equity as a way to further improve it.

## DECLARATIONS

Authors report no conflicts of interest.

## DATA SHARING

NNRD data dictionaries are publicly available at https://digital.nhs.uk/data-and-information/information-standards/governance/latest-activity/standards-and-collections/dapb1595-neonatal-data-set/. NNRD data is accessible via formal application.

## FUNDING

The research is part of a study organised and funded by the AHSN Network (now the NHS Health Innovation Network), the National Institute for Health Research Applied Research Collaboration (NIHR ARC West), and the Heath Foundation. The views expressed in this article are those of the author(s) and not necessarily those of the NIHR or the Department of Health and Social Care.

## Notes

### Competing Interest Statement

The authors have declared no competing interest.

### Clinical Protocols

https://www.medrxiv.org/content/10.64898/2025.12.16.25341898v1

