## Appendix for "Antenatal magnesium sulphate for neuroprotection of preterm babies: data analysis of inequities in treatment in England 2014 to 2024"

**Supplementary materials**

**Appendix 1: Adjustment sets for statistical models**

| **Exposure of interest** | **Outcome** | **Main model 1:**  **Unadjusted** | **Main model 2:**  **TOTAL EFFECTS** | **Supplementary model 3:**  **DIRECT EFFECTS*** |
| --- | --- | --- | --- | --- |
| Maternal age  (≥35 vs <35) | MgSO_4_ | Pre/Post NPP  Interaction between exposure and Pre/Post NPP | *Model 1 +*  IMD  Ethnicity  Region  +date  +clustering on unit | *Model 2 +*  GA  Multiple  Previous pregnancies  Smoking |
| Ethnicity  (white British vs other) | MgSO_4_ | Pre/Post NPP  Interaction between exposure and Pre/Post NPP | *Model 1 +*  No others  *(“No open biasing paths. No adjustment is necessary to estimate the total effect of ethnicity on MgSO4”)*  +date  +clustering on unit | *Model 2 +*  IMD  Maternal age  Previous pregnancies  Region  Smoking |
| IMD  (high vs low deprivation) | MgSO_4_ | Pre/Post NPP  Interaction between exposure and Pre/Post NPP | *Model 1 +*  Ethnicity  Region  +date  +clustering on unit | *Model 2 +*  GA  Delivery mode  Fever in labour  Maternal age  Multiple  Onset of labour  Previous pregnancies  PROM  Smoking |
| Region  (North vs South) | MgSO_4_ | Pre/Post NPP  Interaction between exposure and Pre/Post NPP | *Model 1 +*  Ethnicity  +date  +clustering on unit | *Model 2 +*  GA  IMD  Delivery mode  Fever in labour  Maternal age  Multiple  Onset of labour  Previous pregnancies  PROM  Smoking |

* The main analysis was to estimate the *total effects* of each sociodemographic factor on odds of receiving MgSO_4_. As a supplementary analysis, we also developed adjusted models to estimate the *direct effects* of each sociodemographic factor on the outcome. *Direct effects* represent the component of total effects not mediated by indirect causal pathways(1). For example, the effect of maternal age on receiving MgSO_4_ is likely to act via both direct and indirect pathways; older mothers may have higher odds of treatment as they fall into a higher-risk pregnancy category (a *direct effect* of age), while they may also be more likely to be non-smokers, with higher odds of treatment (an *indirect effect*). In this case, smoking status would be adjusted for in the model that estimates only the *direct effect* of maternal age, but would not be adjusted for in the *total effects* model, which would consider effects across all direct (biological age) and indirect (exposure to smoking) pathways.

1.	Tennant PWG, Murray EJ, Arnold KF, Berrie L, Fox MP, Gadd SC, et al. Use of directed acyclic graphs (DAGs) to identify confounders in applied health research: review and recommendations. International Journal of Epidemiology. 2020;50(2):620-32.

**Appendix 2: Directed Acyclic Graphs for statistical models**

(a) Maternal age *total effects* model

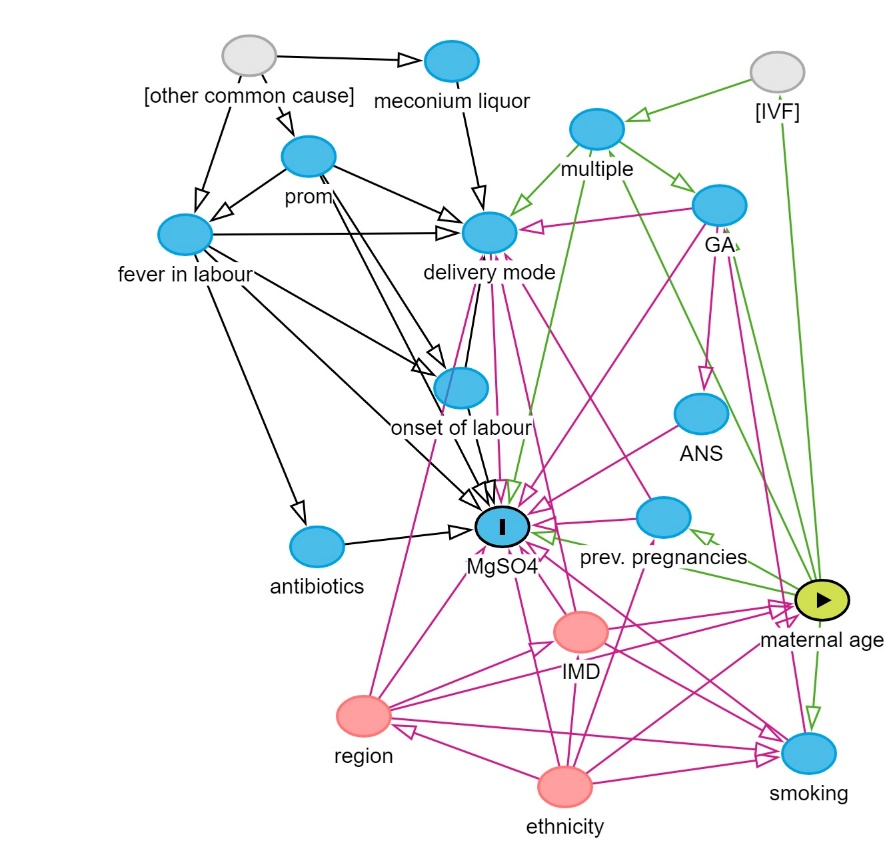

(b) Maternal age *direct effects* model

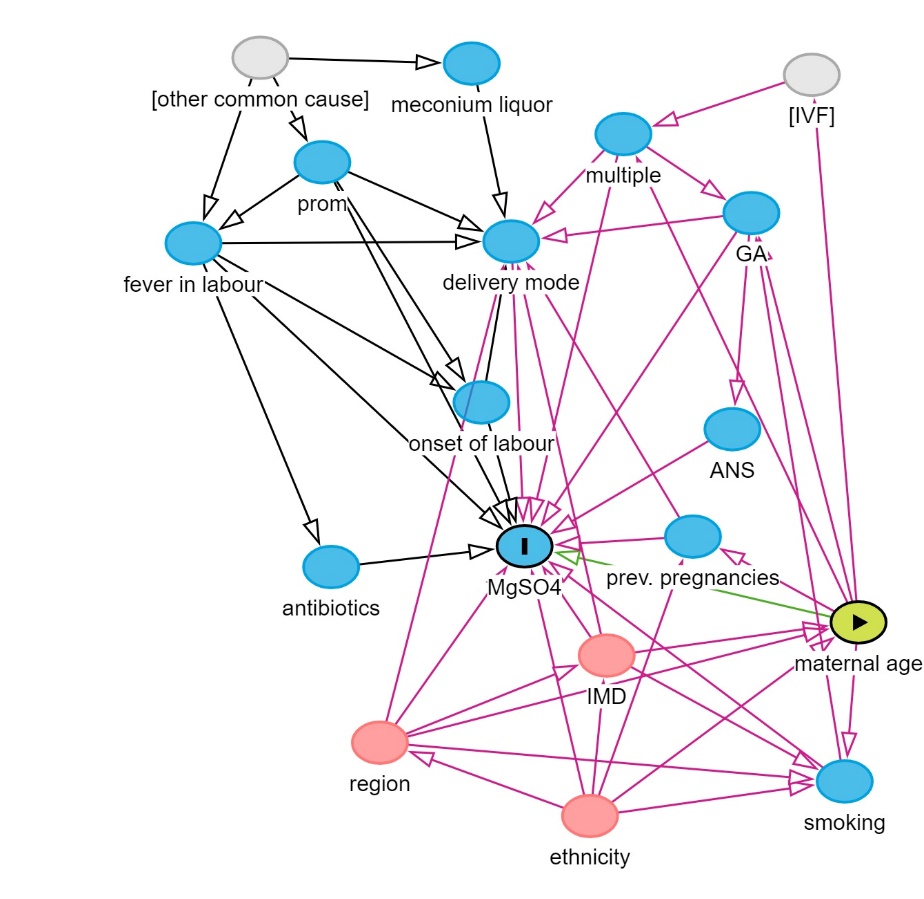

(c) Maternal IMD *total effects* model

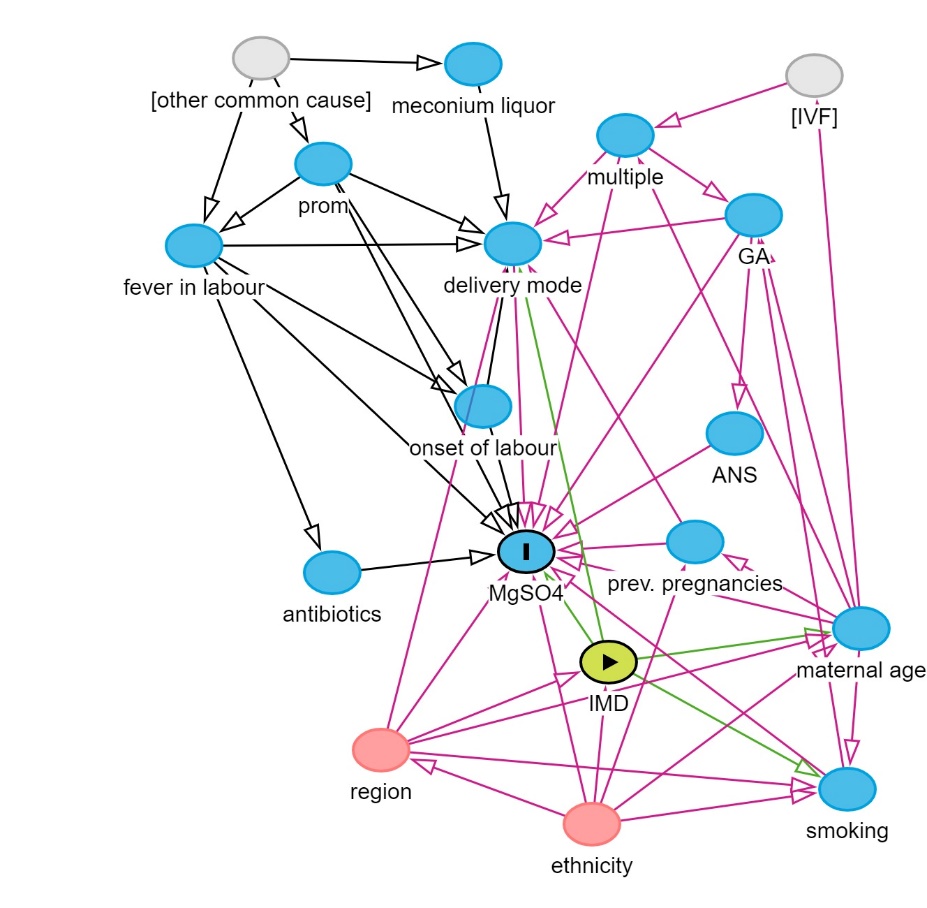

(d) Maternal IMD *direct effects* model

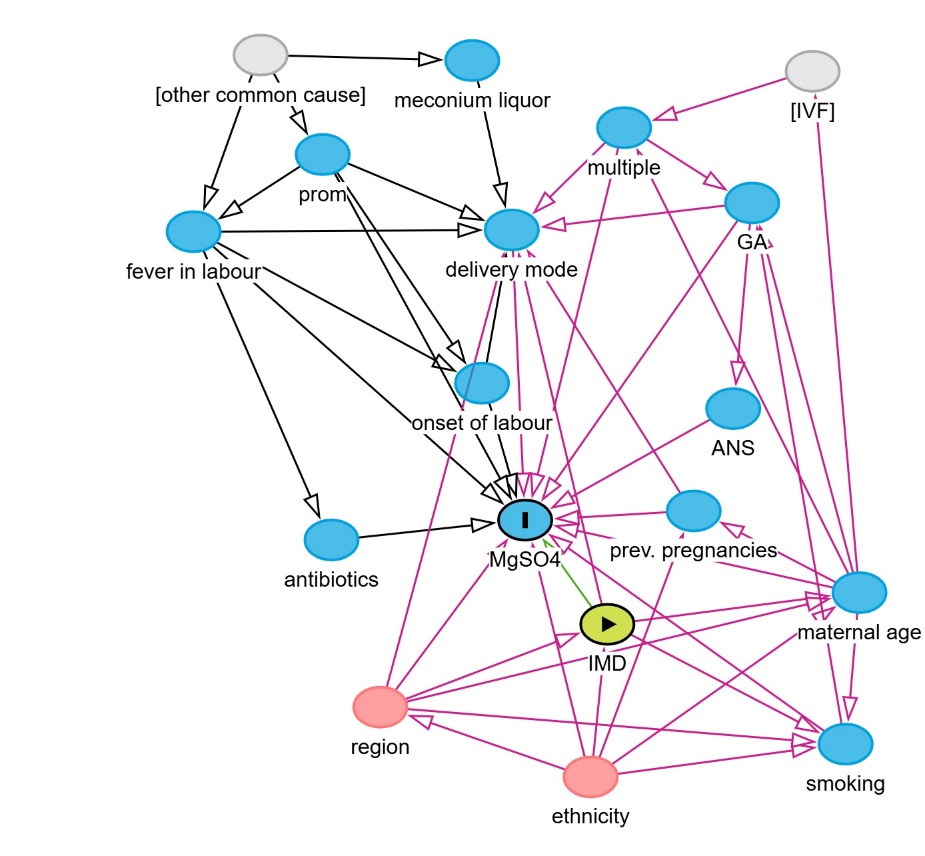

(e) Maternal ethnicity *total effects* model

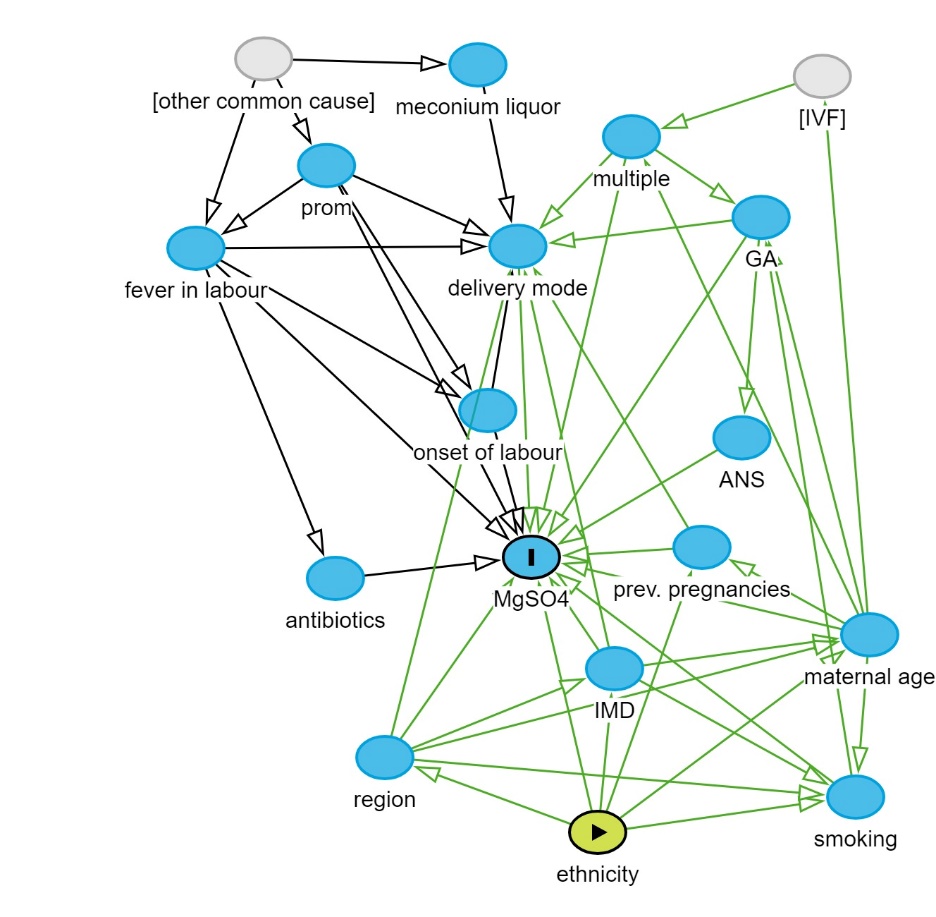

(f) Maternal ethnicity *direct effects* model

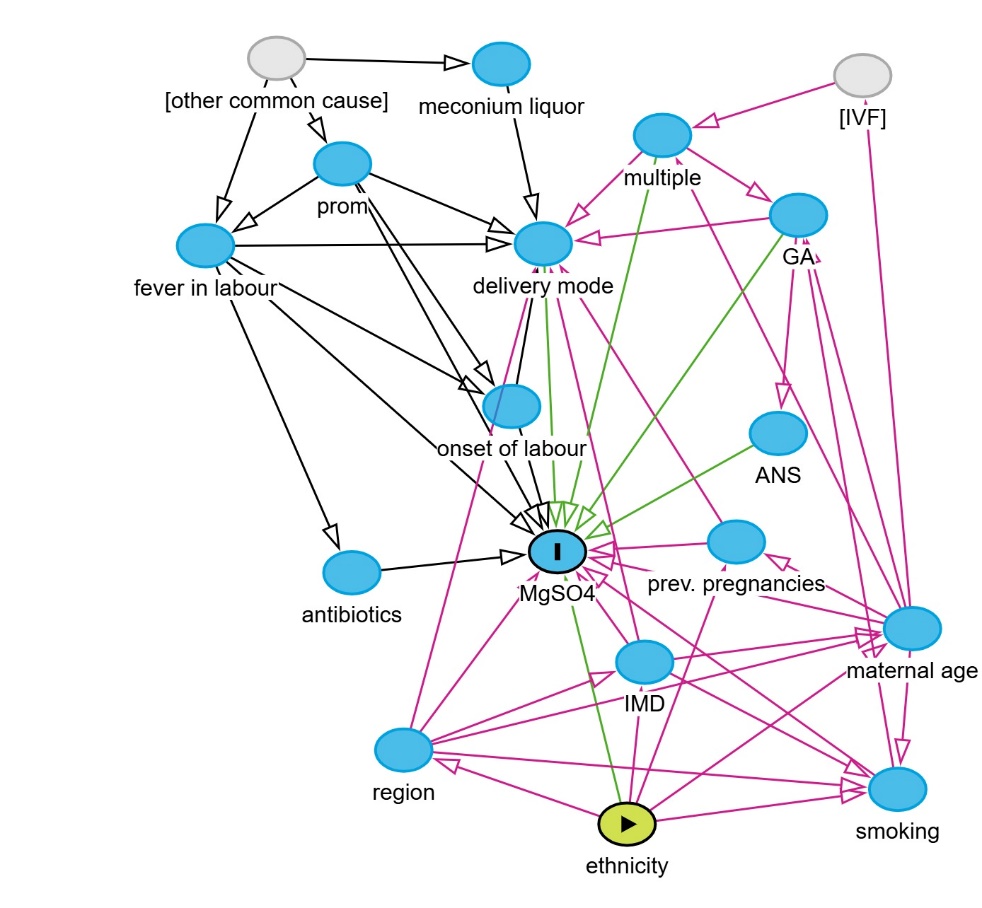

(g) Maternal region *total effects* model

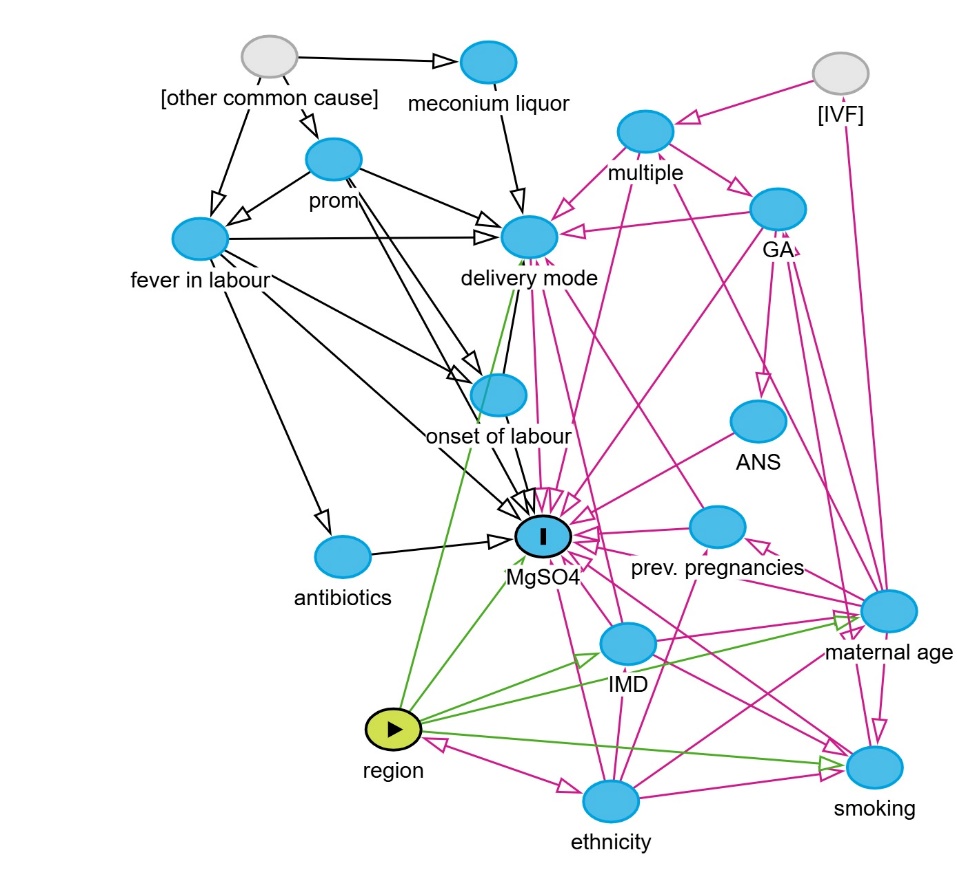

(h) Maternal region *direct effects* model

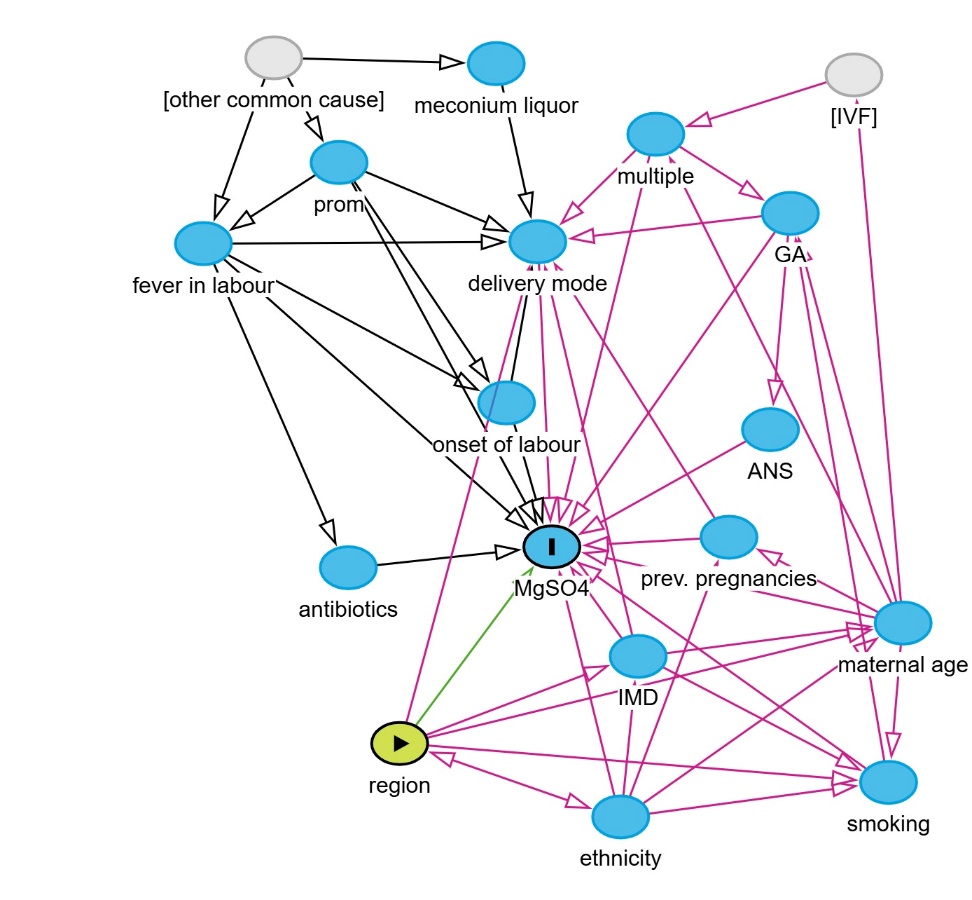

**Appendix 3: Directed Acyclic Graphs key**

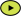
 exposure

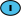
 outcome

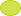
 ancestor of exposure

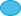
 ancestor of outcome

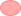
 ancestor of exposure *and* outcome

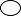
 adjusted variable

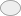
 unobserved (latent)

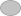
 other variable

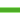
 causal path

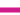
 biasing path

**Appendix 4: Total and Direct effects of sociodemographic factors on odds of receiving antenatal magnesium sulphate for preterm birth in England 2014-2024**

|  | | **Unadjusted model^1^**  **(Odds ratio, 95% CI)** | **Total effects model^1^**  **(Odds ratio, 95% CI)** | **Direct effects model^1^**  **(Odds ratio, 95% CI)** |
| --- | --- | --- | --- | --- |
| **Maternal age (≥35 vs <35 years)** | | | | |
|  | Overall | 1.05 (0.99, 1.11), p=0.127 | 1.01 (0.94, 1.08), p=0.839 | 1.04 (0.97, 1.13), p=0.276 |
|  | p-value for pre/post interaction | p=0.125 | p=0.077 | p=0.255 |
|  | Pre-intervention | 1.09 (1.01, 1.18), p=0.032 | 1.06 (0.97, 1.16), p=0.208 | 1.08 (0.98, 1.19), p=0.120 |
|  | Post-intervention | 0.99 (0.90, 1.09), p=0.828 | 0.93 (0.84, 1.04), p=0.217 | 0.99 (0.88, 1.12), p=0.853 |
| **Ethnicity (white vs non-white)** | | | | |
|  | Overall | 0.88 (0.83, 0.94), p<0.001 | 0.93 (0.87, 1.00), p=0.038 | 0.99 (0.91, 1.07), p=0.785 |
|  | p-value for pre/post interaction | p=0.146 | p=0.235 | p=0.660 |
| **IMD (more vs less deprived)** | | | | |
|  | Overall | 0.88 (0.83, 0.93), p<0.001 | 0.93 (0.86, 0.99), p=0.029 | 1.00 (0.92, 1.08), p=0.915 |
|  | p-value for pre/post interaction | p=0.141 | p=0.022 | p=0.019 |
|  | Pre-intervention | 0.85 (0.79, 0.92), p<0.001 | 0.87 (0.80, 0.95), p=0.002 | 0.93 (0.85, 1.03), p=0.152 |
|  | Post-intervention | 0.93 (0.85, 1.02), p=0.133 | 1.02 (0.92, 1.14), p=0.719 | 1.11 (0.99, 1.26), p=0.080 |
| **Region (North vs South)** | | | | |
|  | Overall | 0.73 (0.69, 0.77), p<0.001 | 0.70 (0.57, 0.85), p<0.001 | 0.70 (0.57, 0.84), p<0.001 |
|  | p-value for pre/post interaction | p<0.001 | p<0.001 | p<0.001 |
|  | Pre-intervention | 0.62 (0.58, 0.66), p<0.001 | 0.60 (0.49, 0.74), p<0.001 | 0.60 (0.49, 0.73), p<0.001 |
|  | Post-intervention | 0.96 (0.88, 1.05), p=0.340 | 0.87 (0.70, 1.08), p=0.201 | 0.88 (0.71, 1.09), p=0.242 |

**Appendix 5: Results of sensitivity analysis excluding mothers with hypertension**

|  | | **Unadjusted model^1^**  **(Odds ratio, 95% CI)** | **Total effects model^1^**  **(Odds ratio, 95% CI)** | **Direct effects model^1^**  **(Odds ratio, 95% CI)** |
| --- | --- | --- | --- | --- |
| **Maternal age (≥35 vs <35 years)** | | | | |
|  | Overall | 1.05 (0.99, 1.11), p=0.135 | 1.00 (0.93, 1.08), p=0.923 | 1.04 (0.96, 1.12), p=0.349 |
|  | p-value for pre/post interaction | p=0.126 | p=0.084 | p=0.281 |
|  | Pre-intervention | 1.09 (1.01, 1.18), p=0.034 | 1.06 (0.96, 1.16), p=0.245 | 1.07 (0.97, 1.19), p=0.161 |
|  | Post-intervention | 0.99 (0.90, 1.09), p=0.827 | 0.93 (0.83, 1.04), p=0.207 | 0.99 (0.87, 1.11), p=0.823 |
| **Ethnicity (white vs non-white)** | | | | |
|  | Overall | 0.87 (0.82, 0.93), p<0.001 | 0.92 (0.86, 0.99), p=0.023 | 0.98 (0.91, 1.06), p=0.636 |
|  | p-value for pre/post interaction | p=0.135 | p=0.207 | p=0.578 |
| **IMD (more vs less deprived)** | | | | |
|  | Overall | 0.88 (0.83, 0.94), p<0.001 | 0.93 (0.87, 1.00), p=0.047 | 1.00 (0.93, 1.09), p=0.911 |
|  | p-value for pre/post interaction | p=0.148 | p=0.022 | p=0.022 |
|  | Pre-intervention | 0.85 (0.79, 0.92), p<0.001 | 0.87 (0.80, 0.96), p=0.003 | 0.94 (0.85, 1.04), p=0.217 |
|  | Post-intervention | 0.93 (0.85, 1.02), p=0.144 | 1.03 (0.92, 1.15), p=0.642 | 1.12 (0.99, 1.27), p=0.067 |
| **Region (North vs South)** | | | | |
|  | Overall | 0.72 (0.68, 0.76), p<0.001 | 0.69 (0.56, 0.84), p<0.001 | 0.69 (0.56, 0.84), p<0.001 |
|  | p-value for pre/post interaction | p<0.001 | p<0.001 | p<0.001 |
|  | Pre-intervention | 0.60 (0.56, 0.65), p<0.001 | 0.59 (0.48, 0.73), p<0.001 | 0.58 (0.48, 0.72), p<0.001 |
|  | Post-intervention | 0.96 (0.88, 1.04), p=0.316 | 0.87 (0.70, 1.08), p=0.206 | 0.89 (0.71, 1.10), p=0.270 |

**Appendix 6: Figures from sensitivity analysis excluding mothers with pregnancy hypertension**

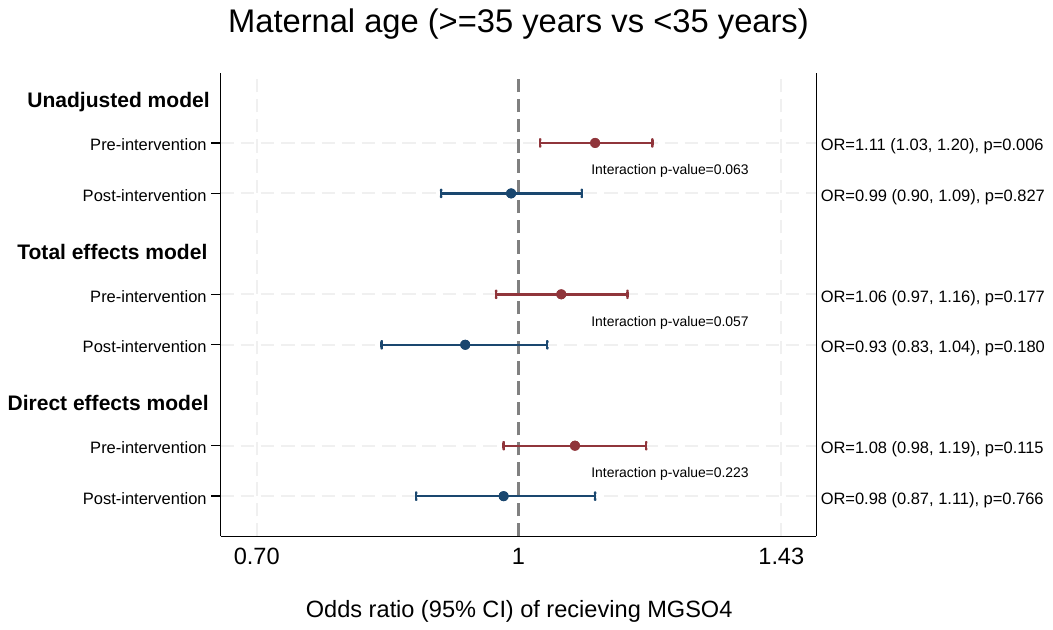

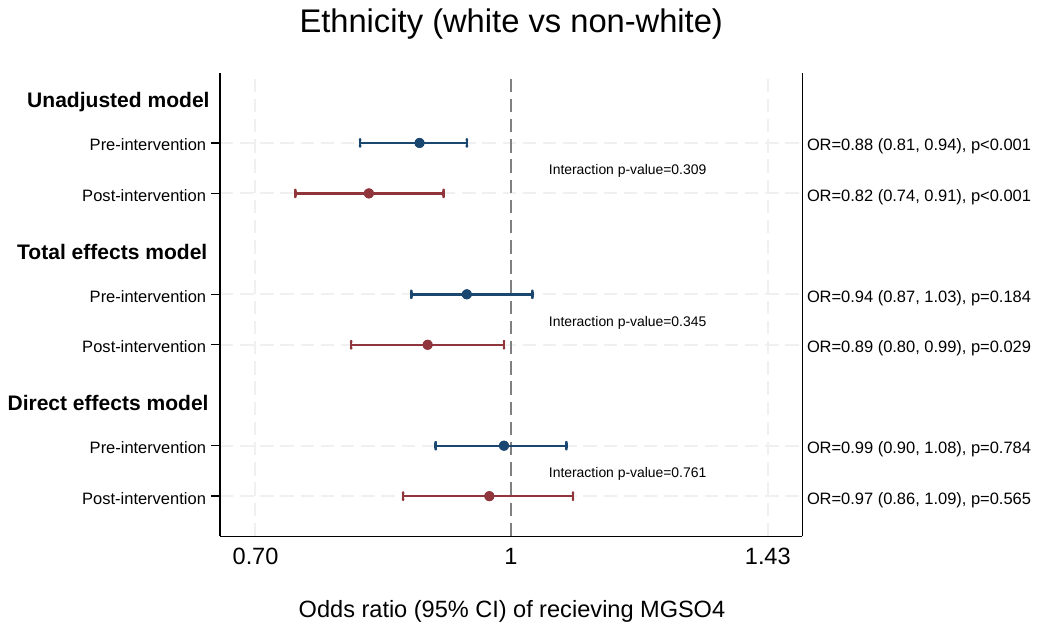

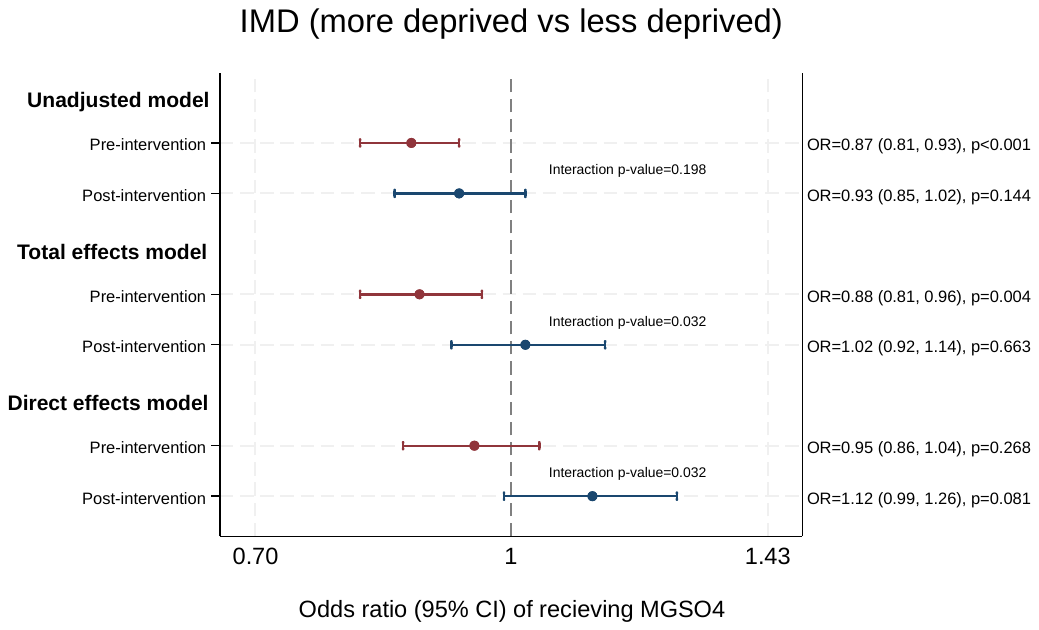

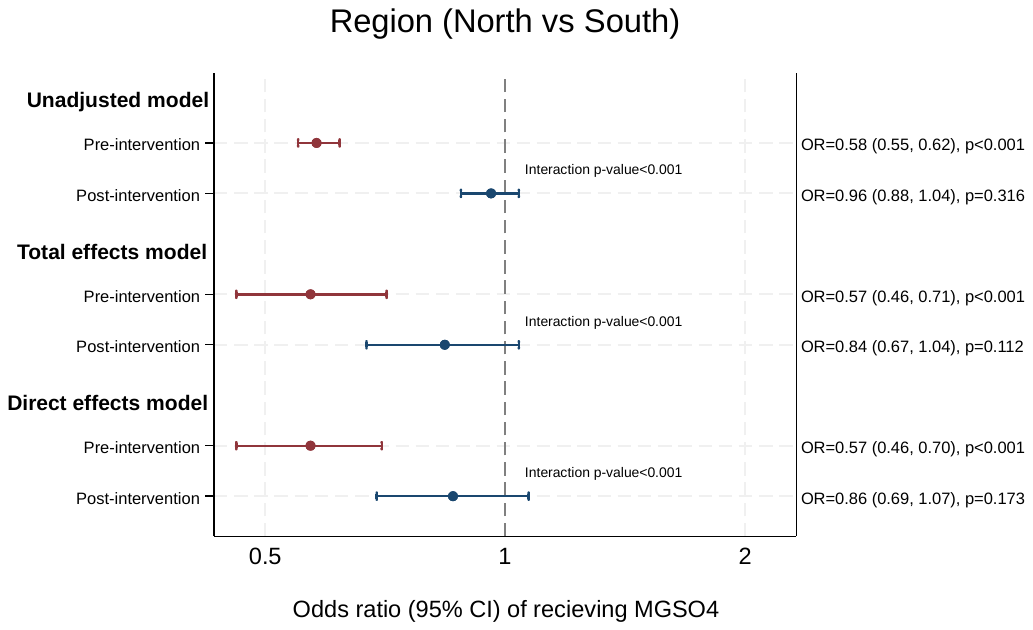
